# CORRELATION OF SERUM ZINC WITH GLEASON SCORE AS A MARKER FOR PROSTATE CANCER AGGRESSIVENESS IN JOS

**DOI:** 10.64898/2026.09.16.26362193

**Authors:** KY Kwarshak, NK Dakum, CG Ofoha, IC Apkayak, VM Ramyil, SI Shuaibu, ZZ Galam

## Abstract

**BACKGROUND:** Prostate cancer is a significant worldwide health issue. The prostate contains the highest zinc concentration among the body’s soft tissues. Zinc inhibits energy production, growth, and proliferation in normal prostate cells while suppressing the malignant potential and angiogenesis that are essential to prostate cancer aggressiveness. The study aimed to determine the correlation of serum zinc with Gleason score as a marker for prostate cancer aggressiveness

**METHODOLOGY:** This was a hospital-based cross-sectional analytical study carried out on forty-two (42) eligible participants with newly diagnosed histologically confirmed prostate adenocarcinoma over one year. The patients were fully evaluated, and 5 mL of venous blood was collected into a plain Vacutainer tube, and a colorimetric test was conducted using Atomic Absorption Spectrometry (AAS). Data for each participant were recorded on a pro forma using KoboCollect software.

The Statistical Package for the Social Sciences (SPSS) version 25 was used for data analysis. All descriptive variables were presented as proportions and numbers. Shapiro–Wilk testing was employed for normality of continuous variables, presented either as median and interquartile range or mean and standard deviation. Spearman’s rank correlation was used to assess the relationship between serum zinc and prostate cancer aggressiveness as measured by the Gleason score. The impact of serum zinc level on the aggressiveness of severe prostate cancer was assessed using a binary regression model, adjusting for age as a confounder in the logistic regression model. A statistically significant P-value was set at ≤ 0.05.

**RESULTS:** The mean age of participants was 67.12 ± 9.68 years, with a range of 45-90 years. High-grade disease (Gleason ≥8) was the most common, representing 57.1% of patients. The median serum zinc concentration was 71.5 µg/dL, with an interquartile range of 47.6–88.5 µg/dL. A statistically significant inverse correlation was observed between serum zinc and Gleason score (Spearman ρ = −0.514, p < 0.001). Reduced zinc levels were significantly associated with higher Gleason scores (Mann–Whitney U = 140.0, p = 0.002). Low serum zinc independently predicted high-grade disease after adjusting for age.

**CONCLUSION:** This study found a statistically significant inverse correlation between serum zinc concentrations and Gleason score in men with prostate cancer. This finding indicates that reduced serum zinc levels are associated with higher Gleason scores.

## BACKGROUND

Prostate cancer (PCa) is a significant health concern among men globally, accounting for the second most common cancer worldwide and the leading cause of death in men, with an estimated 396,792 deaths worldwide.^1,2^ The incidence and mortality rates of prostate cancer rise as age increases, with the average age of diagnosis being 66 years.^2,3^ Disparities in prostate cancer incidence and mortality rates exist between developed and developing nations, partially attributed to sociocultural, genetic, and environmental factors.^4^ In Nigeria, prostate cancer is the most common male cancer with estimated incidence of 46.7%.^5,6^

Zinc is an essential micronutrient found in metalloproteins critical for DNA synthesis, antioxidant function, and immune activity. Among the body’s soft tissues, the prostate contains the highest concentration of zinc.^7^ Compared with normal or hyperplastic prostate cells, malignant prostate cells display consistently low levels of zinc.^8^ Zinc inhibits energy production, growth, and proliferation in normal prostate cells while suppressing the malignant potential and angiogenesis that are essential for prostate cancer aggressiveness.^7,8^ The Gleason grade ranges from 1 (well-differentiated) to 5 (highly undifferentiated). The Gleason Score (GS) represents the sum of two numbers - the Gleason grade of the most dominant and the next most dominant histological pattern. Gleason scores 6 and 7 indicate less aggressive cancer, whereas GS > 7 indicates more aggressive cancer.^9,10^

Despite advances in diagnosis and treatment, understanding the factors contributing to prostate cancer aggressiveness remains incomplete, while its scourge is predicted to continue rising.^11^ While the GS is a widely established, valuable tool for determining the aggressiveness of prostate cancer and potential for disease progression, its drawback remains in inter- and intra-observer variability.^12^ Exploring other biomarkers is essential, as they can increase precision and accuracy when used in addition to the GS. Among the various potential biomarkers, serum zinc levels have emerged as a promising candidate for elucidating prostate cancer aggressiveness and for identifying potential therapeutic agents to mitigate metastasis. Exploring novel biomarkers, such as serum zinc levels, could provide crucial insights into disease aggressiveness, aiding better risk stratification and management strategies. The relationship between serum zinc levels and PCa aggressiveness remains controversial. Some studies have shown a negative correlation, while others have shown a positive correlation.^14–18^ The inconsistency of evidence may be explained by differences in study design and by a lack of accurate or reliable surrogate measures of prostatic zinc in humans. Also, most studies on the correlation between serum zinc levels and GS have been conducted in Western populations, leaving a dearth of evidence in PCa-laden regions such as Nigeria, where presentation is usually late and associated with aggressive cancers. This gap highlights the need for scientifically proven evidence specific to this population. Also, the predictable rise in CaP incidence means that much work needs to be done to unravel possible modifiable predictors of prostate cancer aggressiveness. The study, therefore, aimed to determine the correlation between serum zinc and the Gleason score as a marker of prostate cancer aggressiveness.

## METHODOLOGY

This was a hospital-based cross-sectional prospective analytical study conducted over one year at the Urology Division of Jos University Teaching Hospital (JUTH), Jos, Nigeria.

The study population consisted of men with newly diagnosed histologically confirmed adenocarcinoma of the prostate who presented to the Urology Division during the study period. We recruited all patients with newly diagnosed histologically confirmed adenocarcinoma of the prostate who consented to participate and excluded patients receiving zinc supplementation or medications known to interfere with zinc absorption such as tetracycline, phenytoin, or ethambutol, or patients with medical conditions such as chronic kidney disease and diabetes mellitus that increase zinc excretion, or patients with prior treatment for prostate cancer.

The minimum sample size was calculated using the Leslie–Fischer formula for cross- sectional studies based on an estimated prevalence of prostate cancer of 11% (0.11).^13^ After accounting for a 10% attrition rate, a minimum sample size of 42 participants was obtained. Consecutive sampling was used to recruit eligible participants.

Participants underwent clinical evaluation, including digital rectal examination and serum prostate-specific antigen assessment in the surgical outpatient department. Histological diagnosis was established following digitally guided transrectal prostate biopsy using a spring-driven 18-gauge Tru-Cut biopsy needle. Twelve biopsy cores were obtained from each participant.

A designated histopathologist assessed all the samples and assigned Gleason scores to avoid inter-clinical observer variability. For analysis, Gleason scores were categorised into low/intermediate-grade disease (Gleason ≤7) and high-grade disease (Gleason ≥8). Five millilitres of venous blood were collected under aseptic conditions into plain Vacutainer tubes from eligible participants – histologically diagnosed. Samples were centrifuged within two hours of collection, and serum was stored at −20°C until analysis in the molecular laboratory of the Jos University Teaching Hospital.

Serum zinc concentration was determined using atomic absorption spectrometry with 2-(5- Bromo-2-pyridylazo)-5-[N-propyl-N-(3-sulfopropyl) amino]phenol (5-Bromo-PAPS) reagent. Zinc concentrations ≤84.3 µg/dL were categorised as low, while concentrations >84.3 µg/dL were categorised as normal based on the International Zinc Nutrition Consultative Group reference.^14^

Data analysis was performed using Statistical Package for the Social Sciences (SPSS) version 25.0. Categorical variables were summarised using frequencies and percentages. Age was summarised using mean and standard deviation, while serum zinc and PSA were summarised using median and interquartile range following assessment of normality.

The Mann–Whitney U test was used to compare serum zinc concentrations across Gleason score groups. The association between zinc category and Gleason score group was assessed using Chi-square analysis. Spearman’s rank correlation analysis was used to determine the relationship between serum zinc concentration and Gleason score. Binary logistic regression was performed to determine whether low serum zinc independently predicted high-grade disease, adjusting for age. Statistical significance was set at p < 0.05.

The authors obtained ethical approval from the Jos University Teaching Hospital Research and Ethics Committee. Written informed consent was obtained from all participants prior to enrolment.

## RESULTS

A total of 42 men with histologically confirmed prostate adenocarcinoma were enrolled. The mean age of participants was 67.1 ± 9.7 years, with an age range of 45–90 years. Most participants were between 61 and 70 years of age. More than half of the participants had tertiary education, while retirees constituted the largest occupational group as shown in Table 1.

**Table 1:** Socio-demographic characteristics of respondents with prostate cancer (n = 42)

| Variable | Frequency (n) | Percentage (%) |
| --- | --- | --- |
| **Age Group (years)** |  |  |
| < 50 | 1 | 2.4 |
| 51 – 60 | 6 | 14.3 |
| 61 – 70 | 21 | 50 |
| 71 – 80 | 10 | 23.8 |
| > 80 | 4 | 9.5 |
| **Total** | 42 | 100 |
| **Educational Level** |  |  |
| No education | 5 | 11.9 |
| Primary | 5 | 11.9 |
| Secondary | 10 | 23.8 |
| Tertiary | 22 | 52.4 |
| **Total** | 42 | 100 |
| **Occupation** |  |  |
| Retired | 13 | 31.7 |
| Civil Servant | 9 | 22.0 |
| Businessman | 9 | 22.0 |
| Farmer | 8 | 17 |
| Unemployed | 3 | 7.3 |
| **Total** | 42 | 100 |

The median serum zinc concentration was 71.5 µg/dL, with an interquartile range of 47.6– 88.5 µg/dL. Twenty-nine participants (69%) had zinc levels at or below 84.3µg/dL, indicating a significant prevalence of biochemical zinc deficiency in this group. The Gleason scores were categorised into three: low (Score 6), intermediate (Score 7), and high (Score ≥8). The high-risk group with Gleason scores ≥8 was the most common, accounting for 24 (57.1%) of participants, followed by those with an intermediate score of 7 at 26.2%, and only 7 participants (16.7%) had a low Gleason score of 6, as illustrated in Figure 1.

**Figure 1.** Distribution of Gleason score among participants (n=42)

This study found a statistically significant association between serum zinc levels and Gleason score group (χ^2^ = 27.71, df = 2, p < 0.001). All individuals with Gleason scores of 6 and 7 exhibited normal serum zinc levels, while 82.6% (19 out of 23) of those with Gleason scores of 8 or higher had low serum zinc levels, as shown in Table 2.

**Table 2:** Association Between Serum Zinc Category and Gleason Score Group in participants with prostate cancer (n = 42) cross tabulation.

| Gleason Group | Low Zinc (≤68) | Normal Zinc (>68) | Total |
| --- | --- | --- | --- |
| 6 | 0 | 7 | 7 |
| 7 | 0 | 11 | 11 |
| ≥8 | 19 | 4 | 23 |

| Gleason Group | Low Zinc ($\leq 68$) | Normal Zinc ($>68$) | Total |
| --- | --- | --- | --- |
| Total | 19 | 22 | 42 |

| Test | Value | df | Asymp. Sig. (2-sided) |
| --- | --- | --- | --- |
| Pearson Chi-Square | 27.71 | 2 | <0.001 |

The correlation between serum zinc levels and Gleason score was evaluated in patients diagnosed with prostate adenocarcinoma using the Spearman Rank Correlation test. The results, as shown in Table 3 and Figure 2, demonstrated a statistically significant inverse correlation between serum zinc and Gleason score (ρ = −0.513, p = 0.001, CI: -0.717 to −0.237).

**Table 3:** Non-parametric Spearman Rank Correlation between serum zinc and Gleason score in participants with prostate cancer (n=42)

| Parameter | Value |
| --- | --- |
| Spearman $\rho$ | -0.513** |
| p-value | 0.001 |
| N | 42 |

**\*\* Correlation is significant at the 0.01 level (2-tailed).**

**Figure 2.** scatter plot of serum zinc and Gleason Score in participants with prostate cancer (n=42)

Binary logistic regression analysis was conducted to assess whether low serum zinc levels independently predicted high-grade prostate cancer (Gleason score ≥8), while controlling for age. The findings, as illustrated in Table 4, revealed that low serum zinc was a statistically significant independent predictor of high-grade prostate cancer (B = 2.49, SE =0.88, Wald = 7.84, p = 0.005). Individuals with low serum zinc had approximately 12 times greater odds of having high-grade prostate cancer compared to those with normal zinc levels (OR = 12.06; 95% CI: 2.11–68.93). In the adjusted model, age was not a statistically significant predictor of high-grade disease (p = 0.455).

**Table 4:** Binary logistic regression of serum zinc and Gleason score, adjusting for age in participants with prostate cancer (n=42)

| Variable |  | B | S.E. | Wald | Df | Sig. | Exp(B) | 95%<br>CI Lower | 95%<br>CI Upper |
| --- | --- | --- | --- | --- | --- | --- | --- | --- | --- |
| Low Zinc ($\leq 84.3$ $\mu\text{g/dL}$) | | 2.490 | 0.889 | 7.842 | 1 | 0.005 | 12.063 | 2.111 | 68.926 |
| Age (years) |  | 0.055 | 0.042 | 1.759 | 1 | 0.185 | 1.057 | 0.974 | 1.146 |
| Constant |  | -5.257 | 2.810 | 3.500 | 1 | 0.061 | 0.005 | 0.000 | 1.285 |

## DISCUSSION

This study examined the relationship between serum zinc levels and the aggressiveness of prostate cancer, using the Gleason score as the primary marker of prostate cancer aggressiveness. The key finding was a statistically significant inverse correlation between serum zinc concentration and Gleason score (Spearman ρ = −0.514, p < 0.001). This finding indicates that lower serum zinc levels are associated with higher tumour grades. Low serum zinc also independently predicted high-grade disease, even after adjusting for age.

The majority of participants in this study presented with high-grade disease, with Gleason scores ≥8 accounting for more than half of all cases. This finding is consistent with reports by Wakwe et al.^15^ and Igbokwe et al.^16^ who found a higher proportion of higher Gleason scores among men with prostate cancer in Nigeria. This finding is significant and indicates a pattern of biologically aggressive disease at the time of diagnosis.

In contrast, Margolis et al.^17^ found low Gleason score distribution among prostate cancer patients in the United States, with about 79% having a Gleason score of 7 or less. The high percentage of low-grade tumours in this developed nation suggests that some unexplained genetic and environmental factors are at play.

There was considerable variation in serum zinc levels among the study participants. The median concentration was 71.5µg/dL, with an interquartile range of 47.6 to 88.5 µg/dL. Twenty-nine participants (69%) had zinc deficiency, with levels below 84.3 µg/dL. These findings are similar to other studies by Costello et al.,^18^ Leslie et al.,^11^ and Goel et al.,^19^ which showed low serum zinc levels in patients with prostate cancer compared to healthy adult men or individuals with BPH. These findings underscore the intrinsic role of zinc in inhibiting mitochondrial aconitase, an enzyme that halts oxidative metabolism in the prostate, thereby depriving prostate cells of energy for malignant growth.^20^ Moreover, prostate cancer patients are usually aged in their seventh decade, an age group that is predisposed to malabsorption of nutrients, including zinc, due to intestinal villous atrophy. On the other hand, some studies by Troung-Tran et al.^21^ and Kolonel et al.^22^ have demonstrated higher serum zinc levels in patients with prostate cancer. These apparent discrepancies can be explained by methodological variability. These studies primarily evaluated dietary zinc intake or supplemental zinc exposure through interviews, records and questionnaires rather than directly measured serum or tissue zinc concentrations, which are more reliable surrogate markers. And dietary intake or supplemental zinc does not necessarily reflect biologically available zinc because genetic factors, absorption, systemic transporter activity, and systemic inflammation influence zinc homeostasis.^23,24^

The inverse correlation observed in this study aligns with results from studies across various tissues. Wakwe et al.^15^ in Southern Nigeria demonstrated a strong inverse correlation between plasma zinc and prostate cancer aggressiveness among 220 men with prostate cancer (p<0.001) and determined the significant impact of low zinc in predicting severe prostate cancer (Crude odds ratio: 8.714; p< 0.001). Similarly, Abhishek et al.^25^ found among 102 Indian men with prostate cancer a strong negative Spearman correlation between blood zinc levels and the Gleason score. Further, Cortesi et al.^26^ showed that zinc levels decreased progressively as the Gleason score increased in prostate tissue using X-ray fluorescence analysis among 598 Israeli men with prostate cancer. They found that high-grade tumours had significantly lower zinc concentrations than low-grade tumours, with strong diagnostic power. Likewise, Banas et al.^12^ used prostate tissues from 12 Polish men who had undergone radical prostatectomy and confirmed that tissue zinc levels decreased as Gleason grade increased. These tissue and blood-level findings align with the current serum findings, which suggest that systemic zinc depletion may indicate underlying metabolic changes in the prostate. The biological explanation for this consistent inverse relationship lies in zinc’s unique role in prostate function. Normal prostate cells take up high levels of zinc. High prostatic zinc inhibits mitochondrial aconitase, an enzyme that regulates the Krebs cycle and reduces ATP production, thereby mitigating rapid cancer growth. However, decreased zinc allows aconitase activity to remain unabated, leading to rapidly proliferating prostate cancer cells.^20^ This intrinsic zinc metabolic role in prostate physiology helps explain the inverse Spearman correlation between zinc and prostate cancer aggressiveness observed in this study. In contrast, Igbokwe et al.^16^ in Western Nigeria found a direct relationship between toenail zinc and Gleason score among 41 Nigerian men with prostate cancer (t = 3.463; p = 0.0386). The use of toe-nail for zinc determination has drawbacks. Toenails are exposed to contaminants, including footwear, soil, and dust, which can inevitably affect the true zinc level. Also, toe-nail zinc reflects long-term zinc exposure, explaining the opposite findings. Kolonel et al.^22^ found that high dietary zinc intake was linked to an increased risk of aggressive prostate cancer among older men in Hawaii using a case-control study. Also, the MCC-Spain study by Gutiérrez-González et al.^27^ reported a higher risk of prostate cancer aggressiveness with increased dietary zinc intake. Again, Zhang et al.^28^ demonstrated increased risk of developing aggressive and lethal prostate cancer in US men who have used or are using zinc compared to never-users. These studies examine dietary intake rather than blood or tissue zinc levels and are largely epidemiological in design. Many factors commonly found in the aged, including infection, atrophic intestinal villi, and medications such as phenytoin, can influence dietary zinc absorption.^23,24^ Without adjusting for these factors, it is difficult to conclude that increased dietary zinc increases prostate cancer aggressiveness. Therefore, these differing findings do not undermine the current results but instead reveal methodological or study design differences.

The strength of this study is the adjustment for a confounder – age - to determine the impact of the inverse correlation between zinc and Gleason score.

From a clinical implication, the observation that low serum zinc levels independently predict high-grade disease suggests that it may be a valuable supplementary biomarker for risk assessment. In resource-constrained countries like the Sub-Sahara Africa, where advanced molecular profiling, such as DNA ploidy and tumour angiogenesis, may not be available, serum zinc measurement could provide additional prognostic insights, especially when combined with other established prognostic factors, such as Gleason score. The demonstrated inverse correlation between serum zinc and Gleason scores suggests that serum zinc may not only serve as a biomarker of prostate cancer aggressiveness but also be a potential target for future preventive and therapeutic roles. A further large, multi-centre, prospective cohort study is recommended to establish causality between Serum zinc and Gleason scores, thereby determining potential therapeutic and preventive roles.

## LIMITATION

The cross-sectional design of this study limits the ability of this evidence to establish causality between serum zinc and Gleason score. Also, the digitally guided prostate biopsy used in this study may have affected the evidence.

## CONCLUSION

In conclusion, this study found a statistically significant independent inverse correlation between serum zinc concentrations and Gleason scores in men with prostate cancer. This finding indicates that reduced serum zinc levels are associated with higher Gleason scores. The evidence supports the view that serum zinc is an important biomarker for determining prostate cancer aggressiveness and, could be a potential target for therapeutic and preventive roles.

## Data Availability

All data produced in the present work are contained in the manuscript

## ETHICS STATEMENT

The study involving humans was approved by the Jos University Teaching Hospital Research and Ethics Committee (JUTHREC). The study was conducted in accordance with the Declaration of Helsinki and local legislation and institutional requirements. The participants willingly provided written informed consent to participate after the study’s nature, purpose, and benefits were explained to each participant.

## ACKNOWLEDGMENT

We wish to express our sincere appreciation to Ms. Shangshik Faith Ngwan for her invaluable clerical and administrative support throughout this work. Her dedication, efficiency, and commitment significantly contributed to the successful completion of this project.

